# The risk of future mpox outbreaks among men who have sex with men: a modelling study based on cross-sectional seroprevalence data

**DOI:** 10.1101/2023.08.21.23293147

**Authors:** Marc C. Shamier, Luca M. Zaeck, Hannelore M. Götz, Bruno Vieyra, Babs E. Verstrepen, Koen Wijnans, Matthijs R.A. Welkers, Elske Hoornenborg, Martin E. van Royen, Kai J. Jonas, Marion P.G. Koopmans, Rory D. de Vries, David A.M.C. van de Vijver, Corine H. GeurtsvanKessel

## Abstract

**Background:** In the wake of the 2022-2023 mpox outbreak, crucial knowledge gaps exist regarding orthopoxvirus-specific immunity in risk groups and its impact on future outbreaks.

**Aim:** This study combined cross-sectional seroprevalence studies in two cities in the Netherlands with mathematical modelling to evaluate the risk of future mpox outbreaks among men who have sex with men (MSM).

**Methods:** Serum samples were obtained from 1,065 MSM visiting the Centres for Sexual Health (CSH) in Rotterdam or Amsterdam after the introduction of vaccination and the peak of the Dutch mpox outbreak. For MSM visiting the CSH in Rotterdam, sera were linked to epidemiological and vaccination data. An in-house developed ELISA was used to detect vaccinia virus (VACV)- specific IgG. These observations were combined with literature data on infection dynamics and vaccine effectiveness to inform a stochastic transmission model to estimate the risk on future mpox outbreaks.

**Results:** The seroprevalence of VACV-specific antibodies was 45.4% and 47.1% in Rotterdam and Amsterdam, respectively. Transmission modelling showed that the impact of risk group vaccination on the original outbreak was likely small; however, the number of mpox cases in a future outbreak would be markedly reduced because of vaccination. Simultaneously, the current level of immunity alone may not prevent future outbreaks. Maintaining a short time-to-diagnosis will be a key component of any strategy to prevent new outbreaks.

**Discussion:** Our findings indicate a reduced likelihood of future mpox outbreaks among MSM in the Netherlands under the current conditions, but emphasise the importance of maintaining population immunity, diagnostic capacities, and disease awareness.

## Introduction

Routine smallpox vaccination was discontinued globally in the 1970s following the successful eradication of smallpox. As a consequence, there have been gradual increases in population susceptibility to orthopoxviruses (1), including mpox caused by the monkeypox virus (MPXV). This growing pool of susceptible individuals is thought to have directly contributed to the recent global mpox outbreak with over 85,000 reported cases predominantly among men who have sex with men (MSM) (2, 3). The Dutch 2022-2023 outbreak consisted of over >1,250 reported cases with a peak in July 2022. Of the first 1,000 cases, 99% were males with a mean age of 37, of whom 95% identified as MSM (4).

Prior to the 2022-2023 mpox outbreak, studies conducted in different regions of the world demonstrated significant variations in orthopoxvirus seroprevalence levels. Orthopoxvirus seroprevalence in blood donors was shown to be less than 10% in mpox countries without an animal reservoir of mpox (non-endemic) such as France, the Lao People’s Democratic Republic, and Bolivia (5). In contrast, seroprevalence levels of 51% and 60% were measured in the countries with a mpox animal reservoir such as Côte D’Ivoire and the Democratic Republic of Congo, respectively (6). Although different methodologies were used that limit direct comparisons of seroprevalence rates, these findings underscore high susceptibility on a population level for MPXV infections in non-endemic countries prior to the 2022-2023 outbreak.

A third-generation smallpox vaccine based on the replication-deficient poxvirus modified vaccinia virus Ankara (MVA) (MVA-BN; also known as Imvanex, JYNNEOS, or Imvamune), was rapidly employed in vaccination campaigns during the 2022-2023 mpox outbreak to interrupt MPXV transmission in high-risk populations. We have previously demonstrated that, while a two-dose MVA-BN immunisation series in non-primed individuals induced a cross-reactive immune response against MPXV, levels of MPXV neutralising antibodies were comparatively low (7). Recent studies from Israel (8), the United Kingdom (9), and the United States (10) reported a vaccine effectiveness of MVA-BN against mpox between 36% - 86%, which was comparable to that of the first-generation smallpox vaccine with 58% - 85%(4, 11, 12). Recently, breakthrough infections in previously vaccinated individuals, and re-infections in individuals who had already contracted mpox have been reported (13–17), raising concerns about the longevity of immune responses, and the effectiveness of orthopoxvirus-specific immune responses in preventing novel outbreaks.

In contrast to previous outbreaks of mpox in non-endemic areas (18, 19), the 2022-2023 outbreak exhibited several distinct epidemiological characteristics (3, 20). These included its unprecedented scale, the occurrence of disease mainly among MSM, and sexual contact as a primary mode of transmission (21). A modelling study based on the United Kingdom outbreak highlighted a substantially higher R0 within the MSM sexual network compared to non-sexual household transmissions (21). The outbreak’s deceleration in the second half of 2022 was attributed to the lack of susceptible individuals, either due to vaccination- or infection-induced immune responses, combined with increased awareness, and behavioural changes particularly within the context of sexual interactions (22–24). Despite recognising the importance of population immunity for the prevention of future outbreaks, none of these studies performed immunological assessments and uncertainties persist regarding the current level of immunity among the at-risk population.

To estimate the impact of population immunity on the size and duration of potential future mpox outbreaks, we assessed the seroprevalence of orthopoxvirus-specific antibodies among 1,065 MSM in the two largest cities in the Netherlands after the peak of the 2022-2023 mpox outbreak. The study population comprises MSM presenting at Centres for Sexual Health (CSH), who likely exhibit higher levels of sexual activity than the general Dutch MSM population. Consequently, they are more likely to have been invited for vaccination and/or to have been exposed to the virus, therefore representing the group at highest risk of MPXV infection. The observed seroprevalence levels in combination with published literature data on vaccine effectiveness and infection dynamics were subsequently used in a stochastic transmission model to estimate the magnitude of future mpox outbreaks.

## Methods

### Study population

Centres for Sexual Health (CSH) offer testing for sexually transmitted infections (STI) to those at high risk such as MSM. Mpox testing at CSH was introduced during the early phases of the outbreak in 2022. In the Netherlands, mpox vaccination at Public Health Services started mid-July 2022. Vaccination was offered by CSH to MSM clients who were (current or prospective) HIV pre-exposure prophylaxis (PrEP) users, were HIV infected, or had high risk for STI (defined as notified for STI exposure, having been diagnosed with an STI recently or having multiple sexual contacts). We analysed residual serum samples obtained from these MSM collected at the CSH in Rotterdam and Amsterdam. The sera were collected in September 2022, after the introduction of vaccination and the peak of the mpox outbreak in the Netherlands. The sample set included N=315 sera from Rotterdam and N=750 sera from Amsterdam. Individuals born before 1974 (cessation of smallpox vaccination for the general population in the Netherlands) were inferred to have received childhood smallpox vaccination.

### Ethical statement

Prior to the study, we obtained approval from the Erasmus MC Medical Ethics Review Committee (MEC-2022-0675) granting permission to conduct research using residual materials. The AmsterdamUMC Medical Ethics Review committee granted permission for samples from Amsterdam (W22_428#22.506). A privacy impact assessment was made for combining data from the CSH Rotterdam clients, including vaccination indication and infection status, with their vaccination data. No additional data or samples were collected specifically for this study and all samples were pseudonymised, ensuring that no identifiable data from participants were collected, transferred, or analysed. This study did not involve any direct interaction with participants or any potential harm.

### Detection of VACV-specific IgG antibodies

For the detection of VACV-specific antibodies, an in-house screening ELISA was employed using a VACV Elstree-infected HeLa cell lysate as antigen as described previously (7). Absorbance was measured at 450 nm using an Anthos 2001 microplate reader and corrected for absorbance at 620 nm. Values of optical density measured at a wavelength of 450 nm (OD_450_ values) were obtained with mock-infected cell lysates and subtracted from the OD_450_ value obtained with the VACV-infected cell lysates to determine a net OD_450_ response. A positive control based on a pool of two sera from post-MVA-BN individuals, who had also received childhood smallpox vaccination, was included on every ELISA plate. Based on assay validation, an assay cut-off was defined at OD_450_=0.2 by which a maximum sensitivity (100%) was reached, while maintaining specificity of 89.9% **(Supplementary Figure S1**). This cut-off was chosen to allow for the detection of VACV-specific antibodies in the early stages of infection or shortly after vaccination. A borderline area was identified up to an OD_450_ of 0.35, and the borderline-positive sera are displayed visually distinct from the positive samples in the figures.

### Stochastic model

A mathematical stochastic model was used to model mpox transmission. The model was calibrated to the cumulative number of individuals diagnosed with mpox in the 2022-2023 outbreak in the Netherlands, the seroprevalence at the end of the outbreak, and the number of individuals that were vaccinated (see **Supplementary Figure S2**, **Supplementary Table S1**, and **Supplementary Methods**). The model is seeded by 1-10 individuals that are initially infected with MPXV following an event with high numbers of potential exposures. The model stratified individuals who were not infected based on vaccination status (not vaccinated, historically smallpox vaccinated before 1974, and recently MVA-BN vaccinated during the 2022- 2023 outbreak). Using literature estimates, vaccination is assumed to reduce the risk of infection by 85% (range 75-95%) in historically vaccinated individuals (12) and by 78% (95% confidence interval [CI] 54-89%) in recently vaccinated individuals (8–10). Upon infection, individuals first enter an exposed state in which they are not infectious to others. Individuals become infectious after a serial interval of 8.0 days (95% CI 6.5 – 9.9 days) (25). We assumed that individuals remain infectious to others until they are diagnosed (within 1 to 21 days after symptom onset) (26), after which they will end high risk behavior and will consequently not transmit MPXV to others. At the beginning of the outbreak in 2022, there was no awareness of mpox and diagnostic tests were not available. The outbreak in the Netherlands started between the middle of April and the middle of May 2022. After May 23^rd^, we assumed that awareness increased during the outbreak, resulting in a reduced time between symptom onset and diagnosis in health care, and a reduction in new sexual partners by up to 50% (27, 28). Individuals that recovered from mpox are assumed to not be infectious to others.

To comprehensively assess the impact of varying levels of demographic turnover on the outbreak size, we selected a wide range of turnover rates (1% to 5% per year) while keeping the total population size constant. These percentages were chosen to represent a broad spectrum within which the actual turnover rate likely falls, given the limited availability of specific data regarding demographic turnover within the Dutch MSM population and the ages at which individuals initiate and conclude their sexual activity. This selection also takes into account the historical trend of the crude birth rate in the Netherlands, which decreased from 2.08% to 0.95% between 1960 and 2022 (29).

### Statistical analysis

A chi-square test for equality of two proportions was used to compare the seroprevalence percentage between Amsterdam and Rotterdam. Data were visualised using Prism (v10.0; GraphPad).

## Results

### Study population

Characteristics of the 315 and 750 MSM who visited CSHs in Rotterdam and Amsterdam in September 2022, respectively, are summarised in **Table 1**. Of the 315 MSM visiting the CSH in Rotterdam, the mean age was 34 (IQR [interquartile range] 28 – 42), and 13.6% (43/315) of the subjects were born before 1974 and likely received a childhood first-generation smallpox vaccination. Most of the participants were invited for MVA-BN vaccination due to (current or prospective) PrEP usage (59.4%, 187/315), high-risk behaviour (8.3%, 26/315), or HIV infection (1%, 3/315). At the time of this cross-sectional study, 62/315 (19.7%) subjects had received one dose of the MVA-BN vaccine with a median time between sampling and vaccination of 26 days, and 46/315 (14.6%) subjects had received two doses with a median time between sampling and last dose of 9 days. Five individuals (1.6%, 5/315) among those who visited the Centre for Sexual Health in Rotterdam had tested positive for mpox since May 2022. Of the 750 MSM visiting the Centre for Sexual Health in Amsterdam, the mean age was 32 (IQR 27-40), and 13.3% (100/750) were born before 1974 and presumably received childhood smallpox vaccination. Data on MVA-BN vaccination and MPXV infection were not available for the Amsterdam cohort.

**Table 1.** Characteristics of 1,065 MSM visiting the Centres of Sexual Health in Rotterdam and Amsterdam in September 2022.

|  | Rotterdam<br>N | % | Amsterdam<br>N | % |
| --- | --- | --- | --- | --- |
| **Age** |  |  |  |  |
| <20 | 3 | 1,0% | 12 | 1,6% |
| 20-29 | 101 | 32,1% | 286 | 38,1% |
| 30-39 | 112 | 35,6% | 258 | 34,4% |
| 40-49 | 56 | 17,8% | 102 | 13,6% |
| 50-59 | 28 | 8,9% | 66 | 8,8% |
| 60-69 | 13 | 4,1% | 24 | 3,2% |
| 70-79 | 2 | 0,6% | 2 | 0,3% |
| **Vaccination** |  |  |  |  |
| MVA-BN only | 97 | 30,8% | N/A |  |
| MVA-BN and historic smallpox vaccination* | 28 | 8,9% | N/A |  |
| Historic smallpox vaccination* | 15 | 4,8% | N/A |  |
| Unvaccinated | 175 | 55,6% | N/A |  |
| **Indication for MVA-BN vaccination** |  |  |  |  |
| Not invited | 99 | 31,4% | N/A |  |
| PrEP use or waitlist | 187 | 59,4% | N/A |  |
| PLWH | 3 | 1,0% | N/A |  |
| Recent STI exposure or treatment, or multiple sexual partners | 26 | 8,3% | N/A |  |
| **MVA-BN vaccination** |  |  |  |  |
| 1 dose | 62 |  | N/A |  |
| Time since last dose (mean, IQR) | 26 (18,75 – 33) |  | N/A |  |
| 2 doses | 46 |  | N/A |  |
| Time since last dose (mean, IQR) | 9 (7 – 13,5) |  | N/A |  |
| **PCR-confirmed infections** |  |  |  |  |
| Infected | 5 | 1,6% | N/A |  |

\*Historic smallpox vaccination was inferred for those born prior to cessation of childhood smallpox vaccination in the Netherlands in 1974
MVA-BN, modified vaccinia Ankara – Bavarian Nordic; PrEP, (HIV) pre-exposure prophylaxis; PLWHA, people living with HIV; PCR, polymerase chain reaction; IQR, interquartile range.

### Seroprevalence and serological profiling of orthopoxvirus-specific antibodies

VACV-specific IgG antibodies were detected in 143/315 sera (45.4%) from MSM in Rotterdam. Of these positive sera, 18/143 (12.6% of positives, or 5.7% of total) were borderline positive (OD_450_ between 0.2 and 0.35). VACV-specific IgG antibodies were detected in 353/750 (47.1%) sera from MSM in Amsterdam. Of these, 68/353 (19.3%) were borderline positive (**Figure 1A**). The seroprevalence was lowest in 20-29-year-olds, which comprised most MSM; it was highest in the oldest group of 70-79 (**Figure 1B**). In all groups above 50 years of age, who have likely been historically vaccinated against smallpox, orthopoxvirus-specific antibodies were measured in at least 50% of individuals. Overall, the seroprevalence of orthopoxvirus-specific antibodies among MSM was comparable between Rotterdam and Amsterdam (χ^2^ = 0.257, p = 0.61).

**Figure 1:**
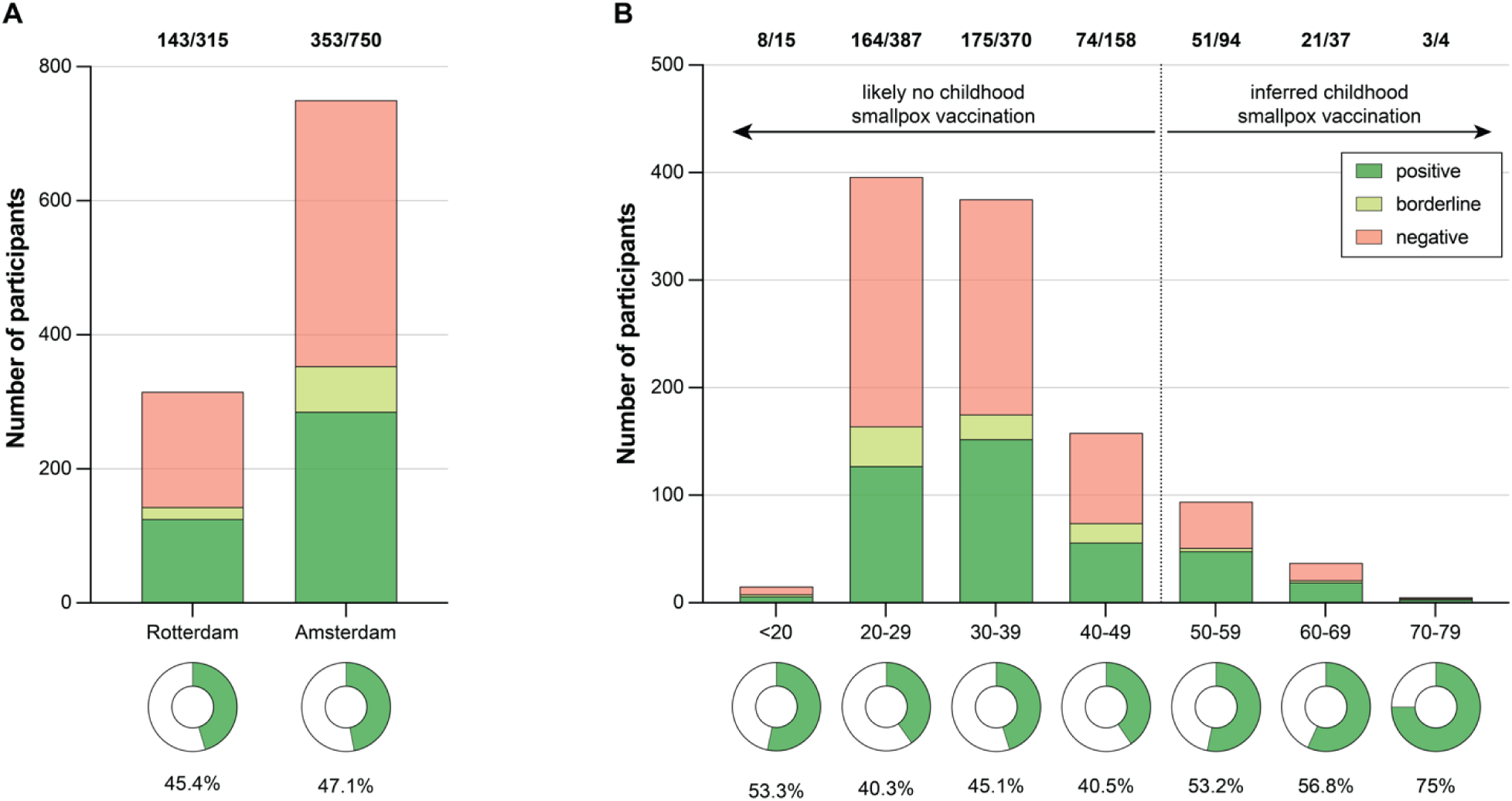
Seroprevalence of orthopoxvirus-specific antibodies among men who have sex with men (MSM) in Rotterdam and Amsterdam. (**A,B**) Detection of VACV-specific IgG in n = 1,065 serum samples from MSM visiting the Centres for Sexual Health in Rotterdam (n = 315) and Amsterdam (n = 750) using an in-house screening ELISA grouped by location (**A**) or age (**B**). Samples were considered as positive with an OD_450_>0.35 (green), as borderline-positive with an OD_450_ between 0.35 and 0.2 (yellow), and as negative with an OD_450_<0.2 (red). Seroprevalence levels were estimated based on the more relaxed cut-off of OD_450_>0.2, including borderline-positive samples, and are shown as donut graphs. Bold numbers above the plots indicate the number of seropositive participants among the respective population. VACV, vaccinia virus.

### Fitting of the stochastic MPXV transmission model

A stochastic MPXV transmission model was generated to estimate the risk of a future mpox outbreak among MSM using our serological observations and available literature data on MPXV infection dynamics and vaccine effectiveness. This model was calibrated to parameters derived from the Dutch 2022-2023 mpox outbreak. The model-generated simulation demonstrated comparability of daily incidence to the real-world data from the Dutch National Institute for Public Health and the Environment (RIVM) (30) of the Dutch 2022-2023 mpox outbreak (**Figure 2A**). Simulation of the 2022-2023 Dutch mpox outbreak using our model yielded a median total cumulative case count of 1,325 (IQR 1,262-1,419) over a duration of approximately 25 weeks. The number of actual reported cases in the Netherlands during the same period was 1,259. Simulating the Dutch mpox outbreak in the absence of a vaccination campaign indicated that risk group vaccination only led to a marginal increase of cumulative cases (1,427, IQR 1321- 1565).

**Figure 2.**
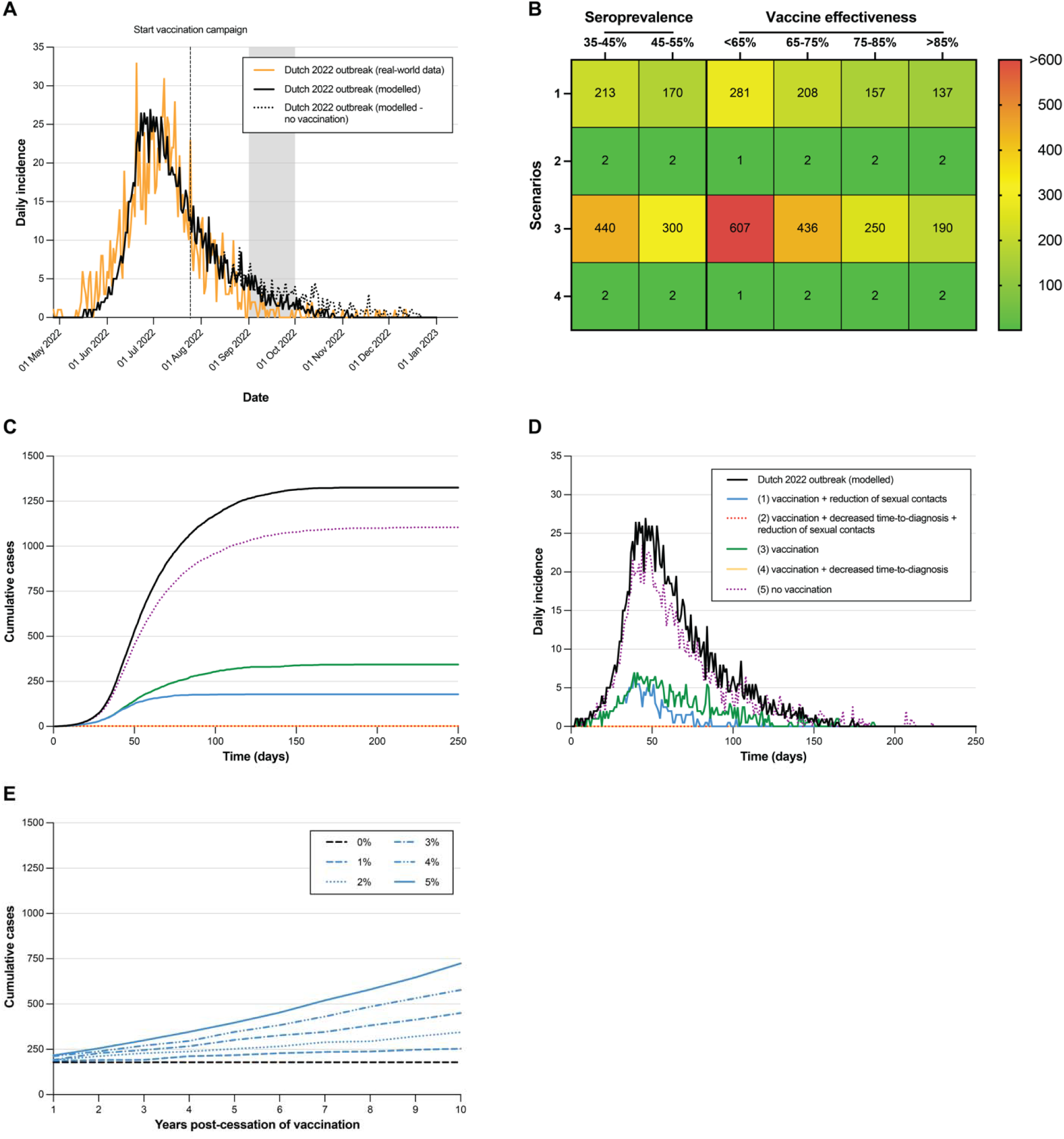
Monkeypox virus (MPXV) transmission model among men who have sex with men (MSM) in the Netherlands. The stochastic transmission model uses the seroprevalence data reported here (range between 35% and 55%; Figure 1) in combination with available literature data on infection dynamics and vaccine effectiveness, and was calibrated to parameters derived from the Dutch 2022-2023 mpox outbreak. (**A**) Comparison of daily incidence from the model-generated simulation (dotted black line) to the real-world epidemiology data (orange line) of the Dutch 2022-2023 mpox outbreak. The start of the vaccination campaign and the sample collection period are indicated. The peak of the curve from real-world epidemiology data was overlayed with the peak of the model-generated curve. (**B**) Sensitivity analysis of the impact of a different seroprevalence or vaccine effectiveness on the cumulative number of mpox cases in a new outbreak. The number in the first column represent the four different modelled scenarios: (1) a partially vaccinated population (seroprevalence 35%-55%) with a reduction of sexual partners within the risk group comparable to the original outbreak, (2) a partially vaccinated population (seroprevalence 35%-55%) with a reduction of sexual partners within the risk group comparable to the original outbreak, and decreased time-to-diagnosis comparable to the end of the outbreak, (3) same as scenario 1 but without a reduction of sexual contacts, and (4) same as scenario 2 but without reduction of sexual contacts (see also Supplementary Methods). (**C,D**) Cumulative cases (**C**) and daily incidence (**D**) over time are shown for all four scenarios (scenario 1: blue line; scenario 2: dotted red line; scenario 3: green line; scenario 4: yellow line) and the model-generated simulation of the Dutch 2022-2023 mpox outbreak (dotted black line). (**E**) Cumulative cases based on the parameters of scenario 1 (seroprevalence between 35%-55%, reduction of sexual partners within the risk group) if an outbreak occurred and no additional vaccination was performed while accounting for population turnover in years after cessation of vaccination. Population turnover was simulated with variable turnover rates between 1% and 5%; 0% represents no turnover and hence maintenance of initial seroprevalence levels. Total population size remained unchanged.

### Vaccination and early diagnosis reduce the size of future outbreaks

A sensitivity analysis was conducted to examine the influence of distinct seroprevalence levels and a varied range of vaccine effectiveness on the stochastic model (**Figure 2B**).

To investigate the impact of immunity conferred by prior infections and vaccinations on a potential future mpox outbreak we conducted several simulations (**Figure 2C,D**). If no vaccination campaign had been started during the 2022-2023 mpox outbreak, a new outbreak would only be slightly reduced in size, with a cumulative case count of 1,105 (IQR, 1,000- 1,206). However, by considering the range of seroprevalence levels between 35% and 55% based on the data reported here, we observed a large reduction in the outbreak size, with a median of 179 cases (IQR 108-265), an average daily incidence of 1.5 cases, and a duration of approximately 17 weeks. This marks an 86.4% reduction in outbreak size compared to our model’s reproduction of the 2022-2023 Dutch outbreak. In this simulation, similar to the outbreak, we assume that the population at risk reduced their number of sexual partners. We separately included a simulation in a vaccinated population, without any change in sexual partners. In this scenario, the total outbreak size was 344 cases (IQR 167-526), with an average daily incidence of 1.8 cases.

Next, we examined the influence of a reduced time-to-diagnosis. At the beginning of the 2022- 2023 outbreak, there was a diagnostic delay, resulting in a delay in case isolation. The duration of infectiousness decreased during the later stages due to reduced time-to-diagnosis and thus earlier isolation. In a simulation involving a partially vaccinated population with a time-to-diagnosis comparable to the later stages of the 2022-2023 outbreak, no outbreak occurred (median case count of 2, IQR 0-6). In this situation, a reduction in risk behaviour did not have any additional impact. Notably, even with lower vaccine effectiveness and seroprevalence ranges, the combination of vaccination and sufficient laboratory testing capacity indicated to be effective in preventing outbreaks.

### In absence of continued vaccination future outbreaks will progressively expand

If vaccination efforts are discontinued, the proportion of susceptible individuals within the MSM population is expected to increase gradually over time due to demographic turnover, as young individuals entering the at-risk group are likely unvaccinated. We modelled the impact of this rising susceptibility, while assuming a stable MSM population size and variable turnover rates ranging from 1% to 5% per year on the basis of scenario 1 (range of seroprevalence levels between 35% and 55%; reduction of sexual contacts by MSM in response to a future outbreak). Our projections indicate that the median size of a potential outbreak in 10 years could vary between 254 cases (IQR 152-363; 1% turnover rate per year) and 725 cases (IQR 523-912; 5% turnover rate per year) if vaccination of at-risk individuals is discontinued (**Figure 2E**).

## Discussion

Here, we show a seroprevalence for orthopoxvirus-specific antibodies of 45.4% and 47.1% among MSM visiting CSH in Amsterdam and Rotterdam, the Netherlands, respectively. Using mathematical modelling, we show that vaccination campaigns had minimal influence on the interruption of the 2022 outbreak, but that these seroprevalence levels will reduce the likelihood and size of future mpox outbreaks. However, to achieve complete prevention of future outbreaks, it is essential to keep the time-to-diagnosis short, similar to the later stages of the 2022-2023 Dutch outbreak. This requires maintained diagnostic capacities, and sustained disease awareness among healthcare professionals and the at-risk groups alike.

We utilised a stochastic approach to model outbreaks of mpox. A simulation of the Dutch 2022- 2023 outbreak demonstrated comparability of our model-generated to the real-world epidemiology data. Assuming no introduction of risk group vaccination during that outbreak we observed only a slight elevation of cumulative case numbers, highlighting that subsidence of the outbreak likely occurred independent of vaccine-induced immunity. Notably, the peak of the outbreak in the Netherlands had already occurred before vaccination campaigns commenced. It was recently suggested that the decline of the outbreak in the Netherlands could have occurred due to infection-induced immunity and behavioural adaptations among highly sexually active MSM (31).

Like any modelling study, the reliability of our findings depends on the underlying assumptions and data used. A strength of our study is that we conducted simulations using the seroprevalence levels measured among MSM in the Netherlands, and combined it with literature data on serial interval (25) and vaccine effectiveness (8–11) that emerged during the outbreak. The practical applicability of mathematical modelling is enhanced by the integration of real-world population immunity data, thereby supporting the formulation of targeted public health responses, including effective vaccination strategies. Furthermore, we included the intended reduction of sexual risk practices among MSM in response to the 2022-2023 outbreak (22, 32). It is however important to recognize that different sexual activity groups have varying contact rates and that the probability of mpox transmission per sexual encounter is unknown. The limited data led us to choose a non-assortative mixing assumption in our model. Predominant transmission within closely related networks of sexually active MSM might affect the outcome of this model; we acknowledge that it might result in an overestimation of future outbreak risks.

These combined factors highlight the intricacies of the actual transmission dynamics and potential biases in the model, urging a careful interpretation of its projections.

Serum samples used for our cross-sectional analysis were collected in September 2022, a period characterised by a rapid decline in the incidence of mpox cases. The vaccination campaign in the Netherlands commenced in July 2022, targeting high risk groups (4). Due to the recent and non-uniform administration of vaccinations to the majority of participants, the timing of this serosurvey might have been premature to capture all seroconversions. To account for the possibility of low antibody titres shortly after vaccination, cut-off values in the VACV IgG ELISA were carefully defined while ensuring high sensitivity and specificity. Considering the potential higher seroprevalence rates in subsequent months, a wider range of seroprevalence levels (up to 55%) was included in the sensitivity analysis of the stochastic model. Even at the upper end of this seroprevalence range, the levels of immunity were insufficient to completely prevent future outbreaks in our model. In addition, vaccination- or infection-induced immunity against mpox is expected to further decline over time, which is indicated by the occurrence of breakthrough infections (13, 14, 33).

To prevent future mpox outbreaks, public health policy should take several factors into consideration. Firstly, population immunity against mpox can decline due to demographic changes within the risk group: as new unexposed and unvaccinated MSM enter the sexually active population, and older individuals who have been vaccinated or recovered from mpox leave the sexually active population, the proportion of susceptible individuals is expected to rise. Assuming turnover rates between 1% and 5%, we simulated median outbreak sizes between 254 cases (IQR 152-363) and 725 cases (IQR 523-912) 10 years from now, respectively, up from 179 cases (IQR 108-265) without population turnover. Secondly, little is known about the longevity of immunity against mpox induced by third-generation smallpox vaccines or previous infection. Accordingly, our simulation of population turnover does not take waning of immune responses into account. As a consequence, our model-generated outbreak sizes 10 years from now likely underestimate total case numbers. In order to assess these changes, longitudinal or repeated cross-sectional studies are needed to monitor immunity levels in at risk populations. Such activities can identify gaps in vaccination coverage across different age groups, enabling focused efforts on younger, previously unvaccinated individuals. Alternatively, it could be necessary to offer booster vaccinations to those previously vaccinated (34). Thirdly, without ongoing outreach and education efforts, there is a concern that MSM may become less aware of the symptoms of mpox over time, potentially leading to increased transmission of the disease.

In conclusion, our study underlines the importance of maintaining mpox-specific immunity in the at-risk population, alongside diagnostic capacities, continuous surveillance, and sustained awareness among healthcare professionals and the at-risk group. These measures are vital for promptly identifying cases and implementing necessary control strategies. In addition, studies are needed to optimize vaccination of persons at risk of spiilover infections in endemic countries. Future research should focus on understanding the longevity of vaccine-induced protection, contributing to a more comprehensive understanding of mpox epidemiology, and facilitating targeted preventive measures.

## Data Availability

Deidentified individual participant data, the analytics code, and other supporting documents will be made available when the study is complete, upon requests made to the corresponding author.

## Acknowledgements

We thank Gini van Rijckevorsel (Public Health Service Amsterdam, Amsterdam, the Netherlands) for initial input in study design and Denise Twisk (Municipal Public Health Service Rotterdam-Rijnmond, Rotterdam, the Netherlands) for help with the collection of epidemiological data.

## Author contributions

Study conceptualisation: MCS, LMZ, HMG, BEV, MRAW, EH, KJJ, MPGK, RDdV, DAMCvdV, CHGvK.

Data curation: MCS, LMZ, HMG, BV, BEV, KW, MRAW, EH, KJJ, RDdV, DAMCvdV, CHGvK.

Formal analysis: MCS, LMZ, RDdV, DAMCvdV, CHGvK.

Investigation: MCS, LMZ, HMG, BEV, KW, MRAW, MPGK, RDdV, DAMCvdV, CHGvK. Methodology: MCS, LMZ, BEV, KW, MRAW, MEvR, MPGK, RDdV, DAMCvdV, CHGvK.

Project administration: MCS, HMG, MPGK, DAMCvdV, CHGvK. Validation: MCS, LMZ, RDdV, DAMCvdV, CHGvK. Visualisation: MCS, LMZ, DAMCvdV, CHGvK.

Supervision: MPGK, RDdV, DAMCvdV, CHGvK. Resources: MPGK, DAMCvdV, CHGvK. Funding acquisition: MPGK, CHGvK.

Writing – original draft: MCS, LMZ, HMG, RDdV, DAMCvdV, CHGvK. Writing – review and editing: All authors.

