## Supplementary Material for "The risk of future mpox outbreaks among men who have sex with men: a modelling study based on cross-sectional seroprevalence data"

[#] shared first authors

* shared last authors

#### Supplementary Figures

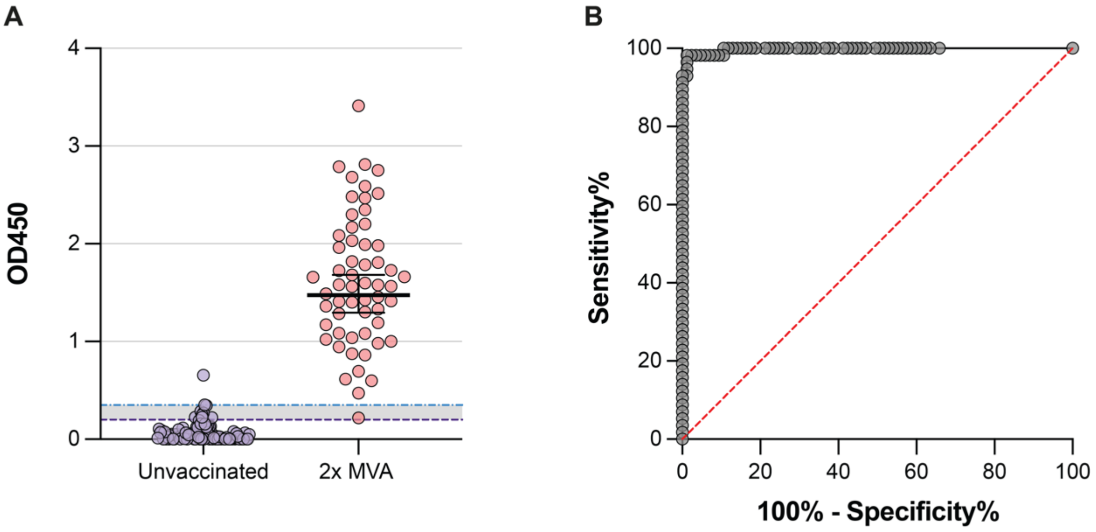


**Supplementary Figure S1. Validation of the VACV-specific IgG screening ELISA.**

(**A**) Distribution of VACV-specific IgG ELISA $OD_{450}$ values in a validation set of 85 sera from orthopoxvirus-naive individuals (expected negative; purple), and a set of 57 sera from double-dose MVA-BN-vaccinated individuals collected 28 days after the second dose (expected positive; red). Samples were interpreted as negative with an $OD_{450}<0.2$, as borderline-positive with an $OD_{450}$ between 0.2 and 0.35 (grey-shaded area), and as positive with an $OD_{450}$ above 0.35. (**B**) An ROC curve was calculated based on the $OD_{450}$ values of the validation set described above (area under the ROC curve = 0.9975 [95% CI 0.9932 – 1.000], $p < 0.0001$).

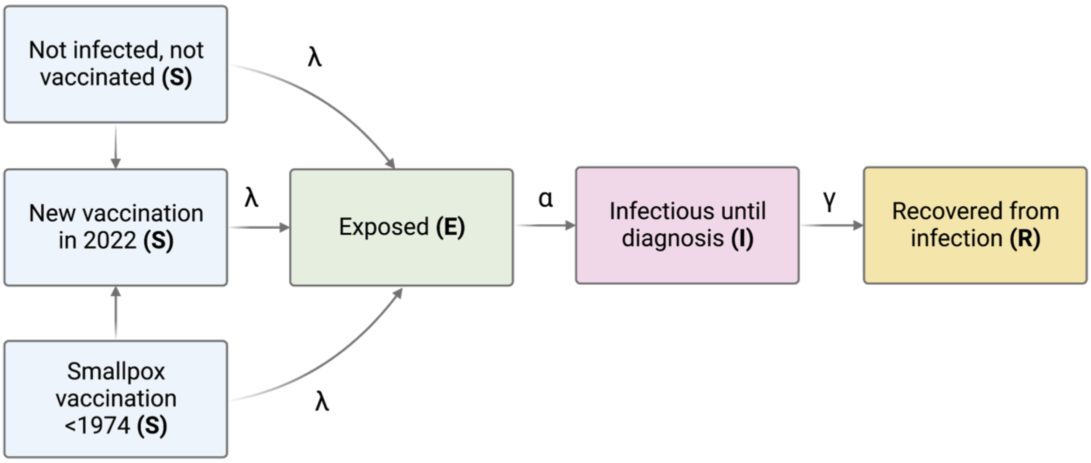


**Supplementary Figure S2. Schematic representation of the mpox transmission model.**

The Susceptible (S) – Exposed (E) – Infectious (I) – Removed (R) model includes three groups of susceptibles (S): naive (not previously infected, unvaccinated), vaccinated with a third-generation smallpox vaccine in 2022, and historically vaccinated before 1974. Susceptibles (S) become exposed to the virus through contact with an infectious individual at a rate of λ. Exposed individuals (E) become infectious (I) at a rate of α, and are removed from the model at a rate of γ, after which they are no longer infectious (R). The arrows in the diagram represent the movement of individuals between compartments. Parameters are further defined in **Supplementary Table 1**.

### Supplementary Tables

##### Supplementary Table 1. Parameters and corresponding values used in the model.

Overview of the definitions of parameters in equations 1-6 (see Supplementary Methods), and the corresponding ranges and references.

| Parameter | Description | Range | Reference |
|---|---|---|---|
| S | MSM at risk of mpox in the Netherlands | 45,000-60,000 | Assumption |
| Vnew | Newly vaccinated | At end of base case scenario 14,000-22,000 | Data |
| Vhist | Historically vaccinated against smallpox | 10%-20% | Data |
| E | Exposed, but not infectious | 0 at start | Assumption |
| I | Infectious | 1-10 at start of outbreak | Assumption |
| R | Recovered from mpox | 0 at start | Assumption |
| λ | Transmissibility of mpox | 0.5-1 | Calibrated |
| Vaccinated | Number of MSM vaccinated during outbreak | 240-360 per day | Calibrated |
| VEnew | Vaccine effectiveness MVA-BN | 78% (95% CI 54%-89%) | [1-4] |
| VEhist | Vaccine effectiveness historical smallpox vaccination | 85% (range 75%-95%) | [5] |
| N | All MSM at risk of mpox | | |
| $pVhist_{(t=0)}$ | Proportion historically vaccinated before 1974 at start of outbreak | 10-20% | Data[6] |
| α | Serial time | 8.0 (95% CI 6.5-9.9 days) | [7] |
| γ | Time from symptom onset to diagnosis or virus clearance | 1-21 days (period 1)<br>4-7 days (period 2) | Calibrated |

### Supplementary Methods

#### Equations used in the stochastic model

The stochastic model based on the Gillespie algorithm[8] can be mathematically described using the following equations:

(1) $S_{(t+1)} = S_{(t)} - \frac{\lambda * S(t) * I}{N} - Vaccinated_{(t)}$

(2) $Vnew_{(t+1)} = Vnew_{(t)} - \frac{\lambda * (1 - VEnew) * Vnew(t) * I}{N}$

(3) $Vhist_{(t+1)} = Vhist_{(t)} - \frac{\lambda * (1 - VEhist) * Vhist(t) * I}{N} - Vaccinated_{(t)} * pVhist_{(t=0)}$

(4) $E_{(t+1)} = E_{(t)} + \frac{\lambda * S(t) * I}{N} + \frac{\lambda * (1 - VEnew) * Vnew(t) * I}{N} + \frac{\lambda * (1 - VEhist) * Vhist(t) * I}{N} - E_{(t)} * \alpha$

(5) $I_{(t+1)} = I_{(t)} + E_{(t)} * \alpha - I_{(t)} * \gamma$

(6) $R_{(t+1)} = R_{(t)} + I_{(t)} * \gamma$

#### Calibration of the model

The model was run 1,000,000 times in MATLAB. A total of 439 simulations were selected, which matched the 2022 outbreak including:

- The cumulative number of mpox cases during the 2022-2023 outbreak (number of 1,200-1,800).
- A deviation of at most 50% in the number of newly reported cases during the first four months of the outbreak (95, 454, 476 and 170 in the first, second, third, and fourth month, respectively)
- The number of newly vaccinated individuals (range 14,000-22,000)
- The seroprevalence of mpox (range 35%-55%)

#### Analysis of the model

For each simulation a unique seeding number was selected, which was subsequently re-used in the analysis of the model to ensure that the same random numbers were chosen in the calibration and in the analysis. In the analysis, we compared four different scenarios:

1. A scenario where a new outbreak occurred under the same conditions as the 2022-2023 outbreak. The seroprevalence in the new outbreak ranged between 35% and 55% (‘vaccination + reduction of sexual contacts’).
2. The same scenario as under 1. In addition, the diagnostic measures implemented during the outbreak continue to be upheld resulting in a reduced time from symptom onset to diagnosis (‘vaccination + decreased time-to-diagnosis + reduction of sexual contacts’).
3. The same scenario as under 1. However, individuals at high risk do not adapt their risk behaviour (‘vaccination’).
4. The same scenario as under 2. However, individuals at high risk do not adapt their risk behaviour (‘vaccination + decreased time-to-diagnosis’).

#### Sensitivity analysis

In a sensitivity analysis, we investigated the impact of the seroprevalence (35%-45% vs 45%-55%) or a different effectiveness of the MVA-BN vaccine (<65%, 65%-75%, 75%-85% and >85%) on the cumulative number of mpox diagnoses during a new potential outbreak (**Figure 2B**).
